# Predicting re-emergence times of dengue epidemics at low reproductive numbers: DENV1 in Rio de Janeiro, 1986-1990

**DOI:** 10.1101/2020.05.02.20074104

**Authors:** Rahul Subramanian, Victoria Romeo-Aznar, Edward Ionides, Claudia T. Codeço, Mercedes Pascual

## Abstract

Predicting arbovirus re-emergence remains challenging in regions with limited off-season transmission and intermittent epidemics. Current mathematical models treat the depletion and replenishment of susceptible (non-immune) hosts as the principal drivers of re-emergence, based on established understanding of highly transmissible childhood diseases with frequent epidemics. We extend an analytical approach to determine the number of ‘skip’ years preceding re-emergence for diseases with continuous seasonal transmission, population growth and under-reporting. Re-emergence times are shown to be highly sensitive to small changes in low *R*_0_ (secondary cases produced from a primary infection in a fully susceptible population). We then fit a stochastic SIR (Susceptible-Infected-Recovered) model to observed case data for the emergence of dengue serotype DENV1 in Rio de Janeiro. This aggregated city-level model substantially over-estimates observed re-emergence times either in terms of skips or outbreak probability under forward simulation. The inability of susceptible depletion and replenishment to explain re-emergence under ‘well-mixed’ conditions at a city-wide scale demonstrates a key limitation of SIR aggregated models including those applied to other arboviruses. The predictive uncertainty and high skip sensitivity to epidemiological parameters suggest a need to investigate the relevant spatial scales of susceptible depletion and the scaling of microscale transmission dynamics to formulate simpler models that apply at coarse resolutions.

## Introduction

Epidemics of arboviruses such as dengue (1), Zika (2, 3), and chikungunya (4) result in substantial global morbidity. Over the past decade, invasions of several arboviruses have triggered large outbreaks in the Western Hemisphere. In Brazil, these invasions include dengue serotype DENV4 in 2012 (5) as well as Zika (2, 6) and chikungunya (7) between 2014-2016. Predicting and understanding the re-emergence of arboviruses after these invasions has important consequences for epidemic preparedness, particularly in regions where climate factors limit mosquito transmission in the off-season. These regions typically exhibit highly intermittent seasonal epidemics, lasting one to three years with long periods of no, or low, reported cases in between, and low mean reproductive numbers (the number of secondary cases arising from each primary case in a completely susceptible population, *R*_0_) (5, 8-10). Several proposed explanations include the depletion of susceptible individuals following initial epidemics (11) and the time required for their replenishment via population growth (12), inter-annual variation in climate (13-17), and antigenic interactions between strains of different serotypes (18-21). These temporal patterns contrast with the recurrent seasonal outbreaks observed in childhood diseases with high reproductive numbers, whose extensive study has provided the basis for our theoretical understanding of SIR (Susceptible-Infected-Recovered) dynamics in infections that confer lifelong or lasting immune protection (22-29).

Statistical models of dengue transmission that take into account climate dependencies can be used to make short-term re-emergence forecasts on the order of 4 months (30) or 16 weeks (15). Many epidemiological models that predict the re-emergence of arboviruses such as Zika (11, 31) on longer time-scales of a year (11) or several decades (31) rely however on compartmental formulations such as SIR-type approaches (11) or Ross-McDonald equations that explicitly incorporate vector transmission (31). Both formulations assume transmission between any two individuals in the population (‘well-mixed’ conditions), typically at aggregated spatial scales. These process-based formulations, for example those recently applied to Zika, represent the acquisition of immunity in the population and its loss via demographic growth and turnover. These models do take into account seasonality of transmission and spatial heterogeneity in the intensity of transmission due to climate at coarse resolutions (at large city, state, or country-level scales). Nevertheless, the replenishment of a well-mixed susceptible population is frequently assumed to be the principal driver determining when the disease will re-emerge given a particular seasonal pattern for *R*_0_ at a particular location(31). Stochasticity can also play an important role in long-term models of re-emergence (31). Variation in reporting rates of arboviruses between locations (32) can add further complexity.

Although childhood diseases with high reproductive numbers display different dynamics from emergent arboviruses (22-26), their compartmental models share a basic SIR structure given the acquisition of long-term immunity after infection. The resulting depletion and replenishment of the susceptible population is known to clearly drive inter-annual variability and re-emergence in the former (25, 27, 28). In particular, recent theory (29) has derived analytical expressions for the number of “skip” years for a measles-like disease in the pre-vaccine era, where “skips” are defined as seasons when transmission occurs but does not cause susceptible depletion. In other words, although the number of infections increases in such seasons, it is not large enough to offset the growth in the susceptible population due to demography. The resulting expressions specifically provide a threshold condition for the number of skips expected following an initial invasion as a function of *R*_0_. Their derivation did not include under-reporting and assumed a closed-population SIR model with ‘school-term’ seasonality, alternating two different rates for low and high transmission.

We examine in this work whether replenishment of susceptible individuals under the typical ‘well-mixed’ assumption explains dengue (DENV1) re-emergence at the whole-city aggregated level. We specifically address the uncertainty inherent in such predictions at the low reproductive numbers characteristic of arboviruses, not previously considered in applications of the analytical approach. To this end, we first extend the threshold derivation to take into account population growth, continuous (sinusoidal) seasonality, and under-reporting of cases. We then fit a stochastic SIR model to observed monthly dengue case counts from the DENV1 invasion in Rio de Janeiro, Brazil from 1986-1988 (8, 10, 33) and numerically predict expected times to re-emergence. We describe high uncertainty in re-emergence times for these seasonal, low transmission regions, and show the insufficiency of susceptible replenishment in a simple SIR model to explain the short periods observed in DENV1 re-emergence. We discuss possible explanations and the need for model formulations that would scale to coarse spatial resolutions.

## Results

We start with the analytical approach for a seasonally forced SIR system with intermittent outbreaks and population turnover, to consider general features of re-emergence at low R_0_. In such a system, the onset of the off-season can bring an end to an initial outbreak, and the replenishment of susceptible individuals due to births and population turnover can be a major determinant of recurrence times. Let S represent the number of susceptible individuals in a population and s_0_, the fraction of the population still susceptible at the end of an initial epidemic, t_0_, when a prediction for the time to the next outbreak will be made. If there are enough susceptible individuals left in the population (i.e. if s_0_ is large), another outbreak will occur in the following year once the on-season resumes. However, if the initial outbreak was very large, s_0_ may be too small, and the outbreak may “skip” one or more years. A skip year is defined as a year in which the susceptible population does not decrease, whether or not infections increase. The smaller the fraction of the susceptible population at the time of prediction (s_0_), the longer it will take for the susceptible population to replenish, and the larger the number of skips that will occur. Previous theory(29) allows prediction of the number of skips that will occur given s_0_. Specifically, it demonstrated that s_0_ must fall below some threshold s_c_ (k) for k skips to occur. An analytical expression was provided for s_c_(k) in terms of the reproductive number and population turnover rate for a closed-population SIR model with school-term seasonality (29). The derivation of the threshold presented in (29) requires the assumption that the transmission rate or reproductive number of the disease is high and that the fraction of the population susceptible at the time of prediction (s_0_) is small.

We extend this approach to take into account population growth and sinusoidal seasonality (which describes the transmission rate of dengue more accurately than a discrete high-low representation). Our derivation does not require assuming that the transmission rate or reproductive number are high or that the fraction of the population susceptible at the time of prediction is small. We follow the criteria developed in (29) (see details in (34)), which essentially consider the sign of the logarithm of the ratio between the respective number of infections at two times, t_0_ and t_n_>t_0_. A positive value indicates that an outbreak will still occur at t_n_; conversely a negative value indicates no outbreak at that time. By setting the logarithm of this ratio to zero, the threshold s_c_ is obtained (See Section 1 of the Supporting Information for details).

The resulting expression for *s*_*c*_*(n)*, the critical fraction of susceptible individuals required at the time of prediction for *n* or more skip seasons to occur, is

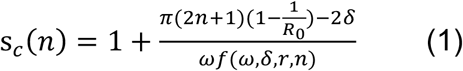

Where 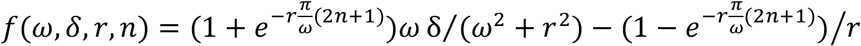, *R*_*0*_ is the annual mean of the reproductive number, δ, the amplitude of seasonal transmission (as infectious contacts per person per day), *ω*, the transmission frequency (in days^-1^) and *r*, the population growth rate (also in days^-1^). The full expression for the seasonal transmission rate is given by *β* (*t*) = *β*_0_(1 + *δsin*(*ωt* + *ϕ*)), where ϕ corresponds to the phase (in radians) and β_0_, to the mean seasonal transmission rate (infectious contacts per person per day). The quantity β_0_ is related to the annual mean reproductive number *R*_*0*_ via the expression *R*_0_ = *β*_0_*>/γ*, where *γ* is the recovery rate (in days^-1^).

Figure 1 illustrates the implications of this formula. As before, t_0_ corresponds to the time of prediction, in practice usually after a large initial epidemic or invasion. Likewise, s_0_ represents the fraction of the population susceptible at the time of prediction. Intuitively, the smaller the fraction of the population susceptible at the time of prediction (s_0_), the longer it will take for the susceptible population to replenish, and the larger the number of skips that will occur. In practice, as we will illustrate below, values of s_0_ can be computed from surveillance data provided one has an estimate of the reporting rate.

**Fig 1.**
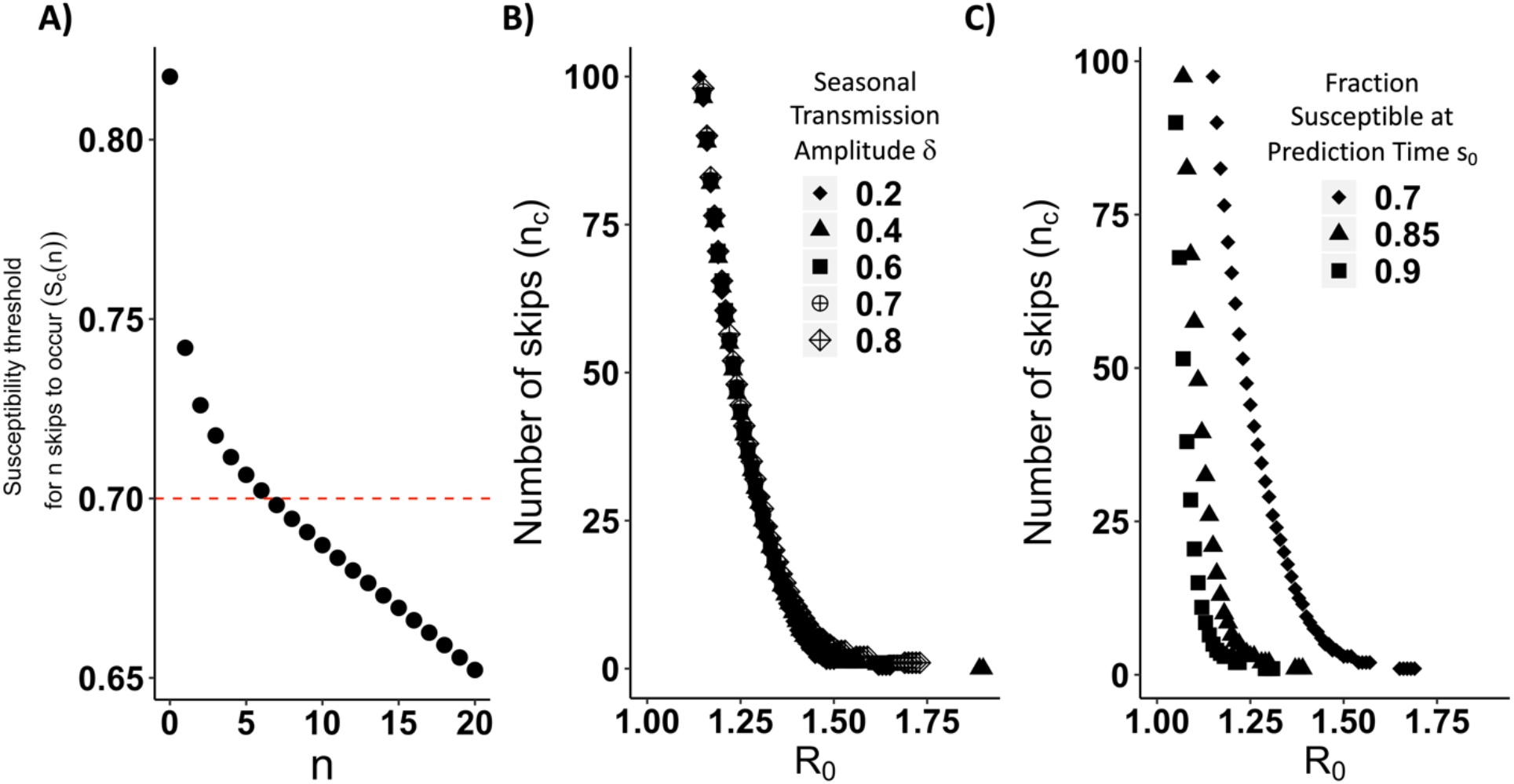
A) Graphical illustration of how the expected number of skips (*n*_*c*_) is calculated. The black dots represent the threshold fraction of the population susceptible at the time of prediction required for *n* skips to occur (*s*_*c*_(*n*)). The plot shows (*s*_*c*_(n)) as a function of *n* (the number of skips) obtained from Equation 1 with seasonality amplitude δ=0.2 (contacts per person per day) and reproductive number R_*0*_=1.4. In this example, the red line represents the fraction of the population susceptible at the time of prediction (*s*_*0*_). If s_0_ is smaller than *s*_*c*_(n), at least *n* skips will occur. To find the expected number of skips (*n*_*c*_), we identify the largest number of skips *n* such that *s*_*0*_ is smaller than the susceptibility threshold required for those skips *s*_*c*_(n). In this example, the red line intersects the *s*_*c*_(*n*) curve between *s*_*c*_(*n*=6) and *s*_*c*_(*n*=7). Therefore, a critical skip number of *n*_*c*_=6 is obtained. **B) and C) The critical skip value n**_**c**_ **as a function of *R***_***0***_ for (B) different values of the amplitude of seasonal transmission *δ* with *s*_0_=0.7 and (C) different values of the fraction of the population susceptible at the time of prediction (*s0)* with *δ*=0.70. In all three panels, the frequency of transmission ω, the population turnover rate μ, and population growth rate r are fixed at respective values ω = (2π/365) day^-1^ corresponding to an annual periodicity, μ= 1/ (74.46*365)) day^-1^ corresponding to an average lifespan of ∼75 years, and r=1.55μ day^-1^ consistent with the growth of the city of Rio de Janeiro. These values were chosen for the purpose of illustration, based on the inverse of the average life expectancy in Brazil in 2012 according to the 2010 census (35), and the interpolation of population estimates for the resident population of the municipality of Rio de Janeiro from the 1991 (36) and 2000 (37) censuses assuming exponential growth.

For *n* skips to occur, the fraction of the population susceptible at the time of prediction (s_0_) must fall below the susceptibility threshold *s*_*c*_*(n)*. Figure 1A shows that the larger the number of skips *n* one is considering, the smaller the threshold *s*_*c*_*(n)* that s_0_ must fall below for at least *n* skips to occur. Let *n*_*c*_ denote the critical skip number corresponding to the number of skips expected at the time of prediction (t_0_). We use the fraction of the population susceptible at the time of prediction (s_0_) and identify the maximum value of *n* for which s_0_ is smaller than *s*_*c*_*(n)*. In the example shown in Fig. 1A, this fraction *s*_*0*_ *=* 0.7 is smaller than *s*_*c*_*(n =* 6) and bigger than *s*_*c*_*(n =* 7), which means *n*_*c*_ = 6. We therefore expect six years of skips followed by re-emergence in the seventh year. Formally, for a given value of *s*_*0*_ at the end of the transmission season, we define the critical skip number *n*_*c*_ as the value of *n* for which *s*_*c*_*(n*_*c*_*) > s*_*0*_ *> s*_*c*_*(n*_*c*_ *+* 1).

With this general approach at hand, we explored the effects of the reproductive number *R*_*0*_, amplitude of seasonal transmission *δ* and fraction of the population susceptible at time of prediction *s*_*0*,_ on the critical number of skips *n*_*c*_ (Figure 1 Panels B and C). Consideration of both the variation of the reproductive number *R*_*0*_ and fraction of the population susceptible at time of prediction *s*_*0*_ is relevant here. Different combinations of transmission rate (β_0_) and duration of the infection (1/ γ) can yield the same *R*_*0*_ but different fractions of the population susceptible at the time of prediction (Supplemental Figure S16). Importantly, Fig. 1 panels B and C show that the time to re-emergence is very sensitive to *R*_*0*_. A singularity is observed as *R*_*0*_ approaches 1 where the expected number of skips goes to infinity. The approach to that singularity can be very steep, meaning that small changes in *R*_*0*_ can result in large increases in the expected reemergence time. The obtained values of *n*_*c*_ are not as sensitive to the amplitude of seasonal transmission (Fig. 1 Panel B) but are sensitive to the fraction of the population susceptible at the time of prediction (Fig. 1 Panel C). The shift of the curve in Fig 1 Panel C for small values of the fraction of the population susceptible at time of prediction *s*_*0*_ means that, for a given *R*_*0*_, more time is required to replenish the susceptible population and therefore to observe a re-emergence.

We next apply this approach to the surveillance data from the 1986 invasion of DENV1 in Rio de Janeiro (Figure 2). The initial DENV1 invasion in Rio de Janeiro is an ideal initial test case for this technique given the lack of widespread prior immunity from to prior dengue epidemics, vaccination campaigns, cross-immunity from other disease outbreaks. Specifically, the 1986 invasion occurred prior to the development of dengue vaccines. The outbreak was the first dengue invasion in the area since the initial eradication of the *Aedes aegypti* mosquito in Brazil in the 1950s (38-41) following a sustained intervention program that began in the 1930s and 1940s in Rio de Janeiro and other cities (39). Cross-immunity from yellow fever vaccination appears to be very limited (42). Given the young age distribution of the population in 1986 (43), most individuals were not alive during the period when mass yellow fever vaccination or prior dengue epidemics occurred.

**Fig 2.**
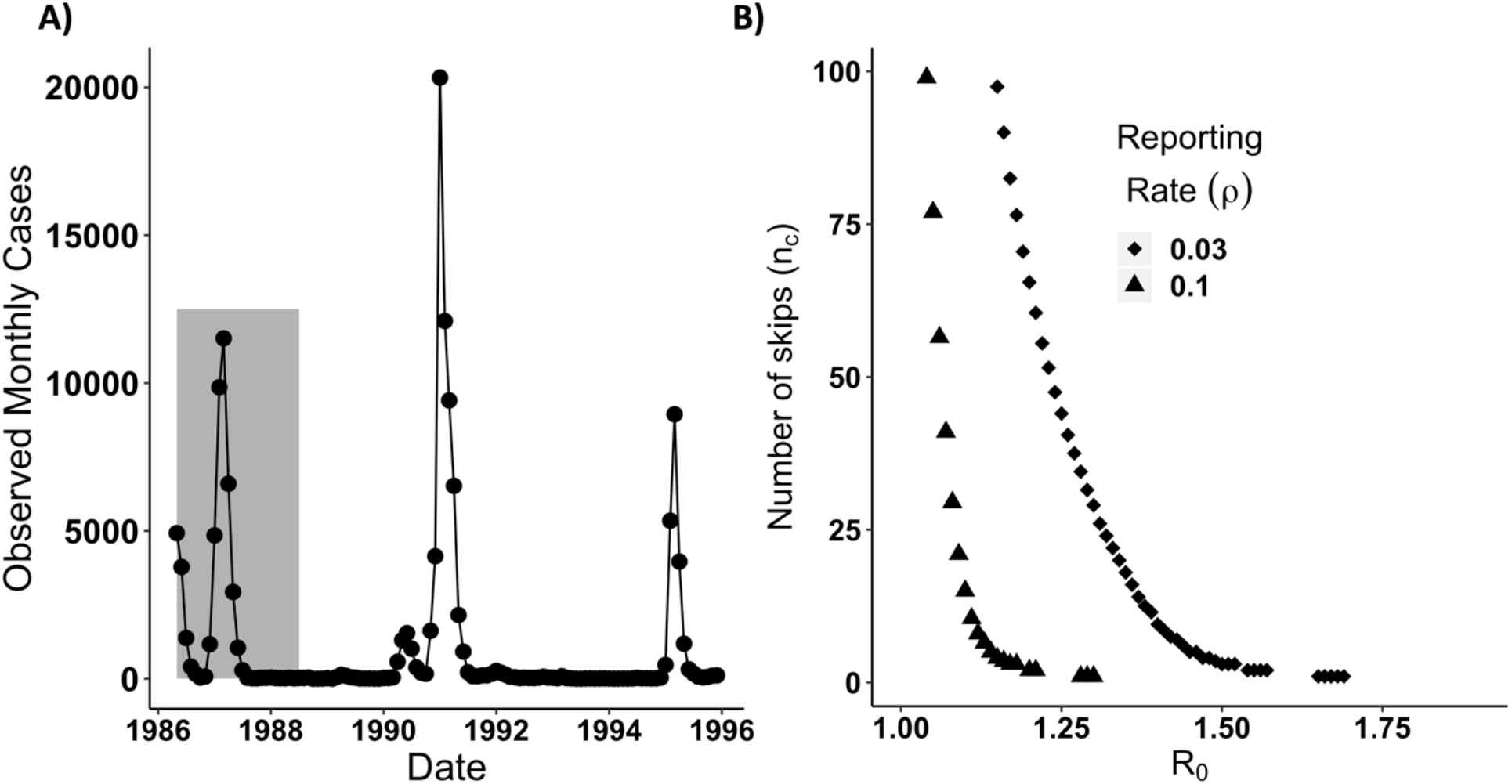
(A) Observed dengue case data. Monthly reported dengue cases in the city of Rio de Janeiro, Brazil from April 1986-1995. The grey shaded region denotes observations that were included in the fitting of the stochastic model from May 1, 1986 to July 1, 1988 inclusive. Serotype DENV1 re-emerged in 1990. DENV2 was first detected in the state of Rio de Janeiro in 1990 but did not become dominant until 1991 (8, 9). Both co-circulated afterwards. We focus on the invasion of DENV1 from 1986-1987 and its initial re-emergence in DENV1 in 1990 using a single serotype transmission model. This allows us to evaluate this transmission model in a region where only one serotype was circulating, where cross-immunity could not easily be invoked to explain the absence or reduction of dengue in a given year. **(B) Deterministic critical number of DENV1 skips *n***_***c***_ **for Rio de Janeiro from September 1988**. Expected number of skips *n*_*c*_ with amplitude of seasonal transmission δ=0.7 and the fraction of the population susceptible after the first DENV1 invasion as of September 1, 1987 (s_0_) calculated from the data (A). We use a reporting rate ρ of 3% when calculating s_0_, consistent with serological estimates from the literature (33). For comparison purposes, we also include the expected number of skips *n*_*c*_ assuming a reporting rate of 10%.

We let our time of prediction t_0_ be equal to September 1, 1987, corresponding to the end of the initial DENV1 invasion (see panel A of Figure 2). In panel B of Figure 2, we evaluate the number of expected skips expected in Rio de Janeiro, *n*_*c*_, on the basis of a range of *R*_*0*_ values from 1.18 to 2.02 from the literature (44, 45). The critical susceptibility threshold for *n* skips to occur (*s*_*c*_*(n)*) is calculated using Equation 1 with an annual seasonality, a population growth rate interpolated from the census (see Materials and Methods section), and δ =0.7 (44). The fraction of the population susceptible at the time of the prediction (*s*_*0*_) is estimated as the difference between the total population N_0_ (total population N at (t_0_=Sep. 1987)) and the total number of people infected between the start of the invasion and the time of prediction (September 1, 1987). The total number of infected people during the outbreak is computed by summing the ratio between the observed monthly cases and the reporting rate for DENV1 in the city. Literature estimates from serology during the DENV1 invasion in Rio de Janeiro indicate a reporting rate of around 3% (33) which we use and fix for this analysis. For comparison purposes, we also include the number of skips expected under a higher reporting rate of 10%. These curves show that the expected re-emergence could be very sensitive to small variation in *R*_*0*_ and ρ, two quantities that are difficult to estimate with precision in the absence of serology. In particular, assuming a reporting rate of 3%, a reproductive number of 1.2 with 20% uncertainty can yield large changes in the expected re-emergence time. We highlight the potential sensitivity of the expected number of skips to the reporting rate as well to illustrate the importance of uncertainty in this parameter in cities or epidemics where its value is unknown.

### Replenishment of Susceptible Individuals is Insufficient to Explain Re-Emergence

To obtain more precise bounds for the reporting rate and *R*_0_ and to determine if the depletion and replenishment of susceptible individuals could explain the rapid re-emergence of dengue in Rio de Janeiro, we fit a stochastic aggregate SIR model to case data from the DENV1 invasion from 1986-1988. The stochastic SIR model assumes that the underlying deterministic transmission rate varies seasonally as a sinusoidal function with annual mean β_0_, seasonal transmission amplitude δ, frequency ω (equal to 2π/365), and phase ϕ. The model takes into account demographic stochasticity, environmental stochasticity in the transmission rate, and measurement error due to under-reporting and variation in reporting of cases (See Materials and Methods and the Supporting Information). Panels A,B, and C of Figure 3 show the likelihood profile of the annual mean transmission rate, β_0_, the amplitude of seasonal transmission δ, and the reporting rate ρ, respectively. In particular, our estimate of the reporting rate matches that from serology in the literature (Panel C).

**Fig 3.**
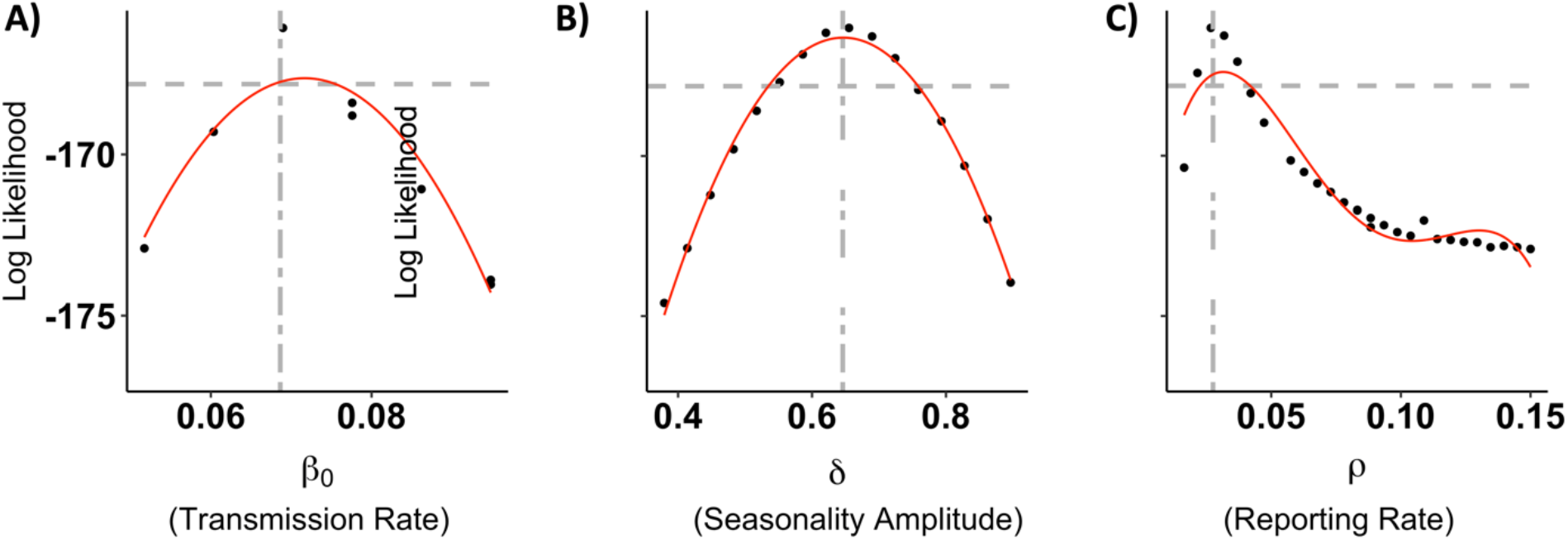
A-C) Selected parameter profiles for the stochastic model. Profiles of the mean annual transmission rate β_0_ (A), seasonal transmission amplitude δ (B), and reporting rate ρ (C). The red curve is a polynomial fit to the subset of the profile points shown on the figure. The single dashed grey horizontal line represents the likelihood value 2 log likelihood units below the maximum likelihood estimate. This line provides an estimate of confidence intervals for the given parameter. The grey vertical line denotes the parameter value of the maximum likelihood estimate. The maximum likelihood estimate for the reporting rate in panel C is very close to the literature value obtained from serology (approximately 3 percent). (33).

Overall, the model is able to capture key dynamics of the DENV1 invasion including the two peaks of incidence in 1986 and 1987 and the subsequent reduction of transmission in 1988. This is shown by comparing the trajectories for an ensemble of simulations with the fitted model to the observed values of cases (Fig. 4). Estimated values for the transmission rate indicate a low value for *R*_0_ (Figure 4 Panel C). Both of these conclusions generally hold even if one takes into account uncertainty in parameter estimates by examining all parameter combinations with log likelihoods within 2 log likelihood units of the maximum likelihood estimate (the grey region in Figure 4 Panel C as well as Supplemental Figure S1), although some parameter combinations (not the maximum likelihood estimate) have substantial process noise (Supplemental Figure S1).

**Fig 4.**
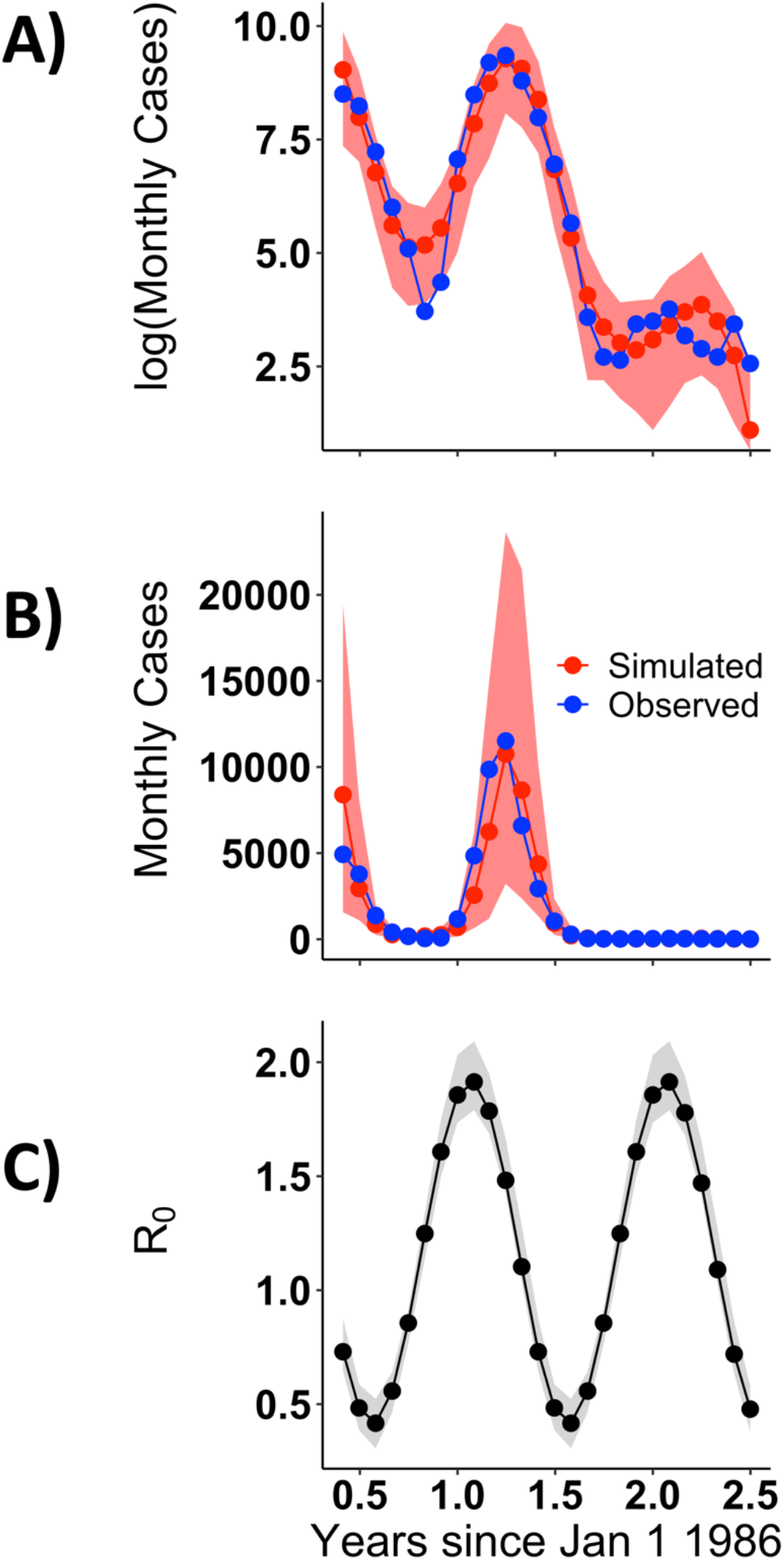
A-B) Comparison of simulated values with the fitted model and observed data on a log (A) and regular (B) scale. Observed monthly cases from April 1986 to June 1988 are shown in blue. Median values from 100 simulations with the maximum likelihood parameter combination are shown in red. The shaded red region denotes the 2.5% and 97.5%th quantile boundaries from those simulations. **C) Estimates for *R***_**0**_***(t)***. The black line denotes the trajectory of *R*_0_*(t)* for the maximum likelihood estimate. The shaded grey region represents the 2.5% and 97.5%th quantile boundaries for trajectories from all parameter combinations within 2 log likelihood units of the maximum likelihood estimate. Each parameter combination has only one seasonal trajectory for *R*_0_*(t)* since *R*_0_*(t)* is a deterministic quantity. *R*_0_*(t)* for all parameter estimates ranges from 1.79-2.09 in the on season to 0.31-0.52 in the off-season.

We now apply the obtained parameter estimates from the fitted model to address the expected re-emergence time on the basis of, first, the analytical expression for the skip calculation (Equation 1), and then the stochastic simulations of the fitted model. The parameter estimates used here are those for the reporting rate ρ, the reproductive number *R*_0_, and the amplitude of seasonal transmission δ from all combinations within 2 log likelihood units of the MLE. The expected number of skips following the DENV1 invasion in 1986-1988 is considerably higher than the observed 2 years. Depending on the parameter combination used, we obtain anywhere from 27 to 100 skips (Panel A of Figure 5). Even the fastest estimated return from the skip analysis (27 years) is much slower than the observed re-emergence time.

**Fig 5.**
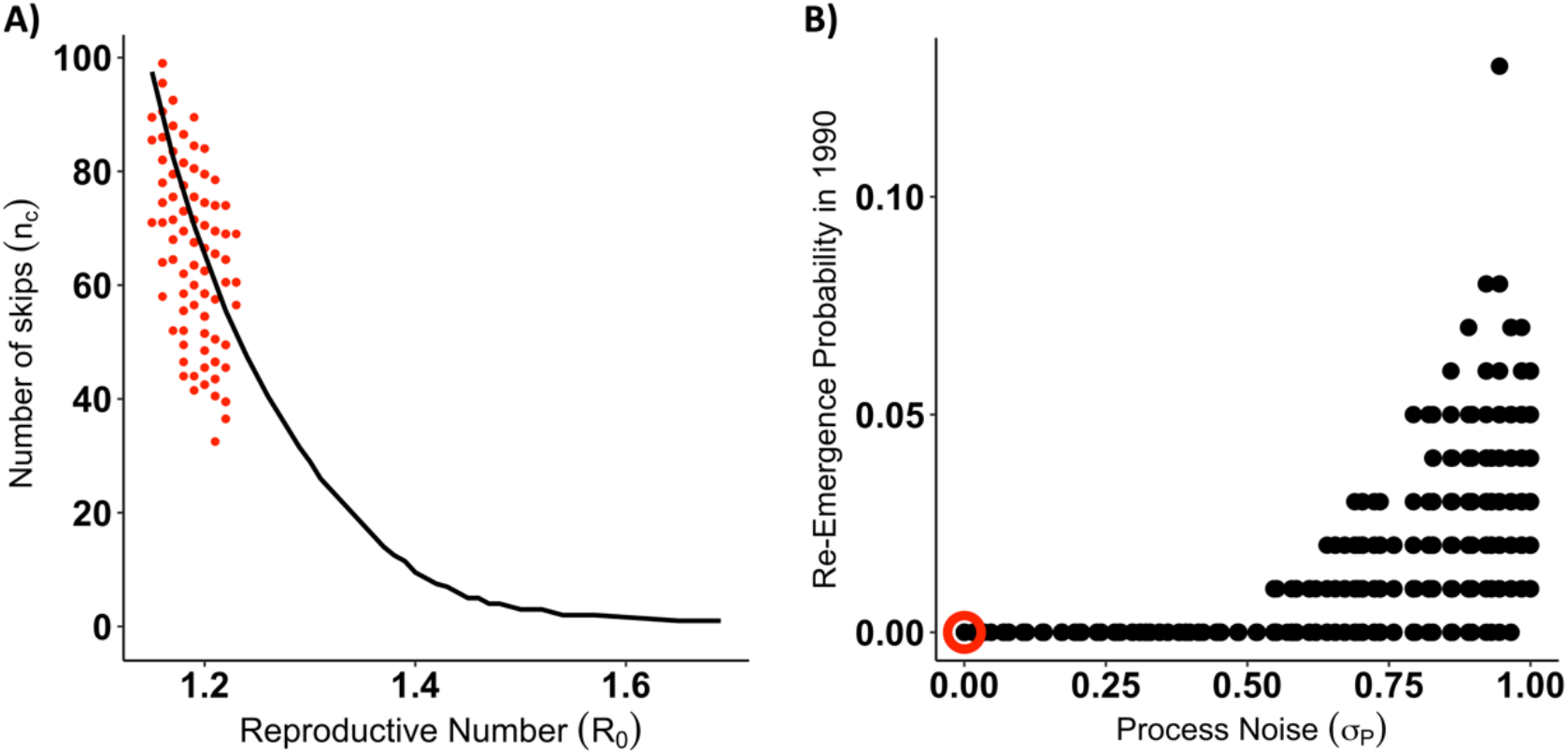
A) Expected number of skips (n_c_) calculated using parameters obtained from the fitted stochastic model. The open circles show the expected number of skips n_c_ from Equation 1 using parameters and the fraction of the population susceptible after the initial DENV1 invasion (s_0_) estimated from the fitted stochastic model. Each circle corresponds to one parameter combination, and we included here all parameter combinations for the fitted model with a seasonal transmission amplitude (δ) of 0.7 (contacts per person per day) and a likelihood value within two log-likelihood units of the maximum likelihood estimate (MLE). See Figure S15 for expected skips from parameter combinations with different values of δ, and Figure S10 for parameter combinations from the profile of the recovery rate, γ. For comparison purposes, the black line shows the expected number of skips for the deterministic skip calculation from panel B of Figure 2 with the reporting rate ρ fixed at the literature value of 3%. **B) Probability of epidemic in 1990 under forward stochastic simulation of fitted model**. The fitted stochastic model was simulated forward in time from 1986-1990 with population growth. A pulse of 20 infected individuals were assumed to arrive each day in January 1990. Each parameter combination within 2 log likelihood units of the maximum likelihood estimate was simulated 100 times. The re-emergence probability was calculated by determining the number of simulations in which the susceptible population decreased in 1990. The plot shows re-emergence probability as a function of the process noise intensity σ_P_. Each point represents a single parameter combination. The maximum likelihood estimate parameter combination is circled in red.

Forward simulation of the stochastic model likewise does not predict the rapid re-emergence of DENV1 (Panel B of Figure 5). Under a pulse of 20 infected individuals arriving per day, there was a low probability of re-emergence for parameter combinations with low process noise (Panel B of Figure 5). Only parameter combinations with high amounts of process noise (which have limited predictive value) had a non-zero emergence probability. We consider alternate pulse rates in Supplemental Figure S14. Re-emergence probabilities under forward simulation of the stochastic model thus corroborated the deterministic skip findings. The depletion of susceptible individuals from 1986-1988 and their replenishment via population growth from 1989-1990 under an aggregate SIR model was unable to explain the rapid re-emergence of DENV1 in 1990.

### Sensitivity Analysis

To examine the robustness of our findings to adding an incubation period or altering the form of seasonality, we conducted a sensitivity analysis by considering both SIR and SEIR models with spline seasonality. The results are presented and discussed in the Supporting Information and show that our conclusions remain unchanged. (See the Supporting Information including Supplemental Figures S2-S7 and Supplemental Tables ST2 and ST3).

### Comparison with Vector Model and literature R_0_

The fitted stochastic SIR model uses a cosine function as a simplification to represent the seasonal forcing that would be created by climate variation (temperature (46)) via the changes in infected mosquitos. To evaluate whether this simplification is realistic, we take two approaches. The first one compares the mean seasonal R_0_ resulting from our model to values of this reproductive number directly estimated from time series data in the literature for DENV1 and DENV4 in Rio de Janeiro from 2010-2016. There is a close match between these very different ways to estimate R_0_, and in particular the shape of the seasonality produced by our model is realistic (Supplemental Figure S18).

The second approach considers a simple temperature-driven vector model. To this end, we initially show that the seasonal variation in temperature in Rio de Janeiro can be approximated via a cosine function (Panel A of Supplemental Figure S19 and use this approximation to drive a transmission rate that includes the vector explicitly.

To obtain an expression for the seasonal transmission rate we consider an explicit mosquito model with compartments for infectious and susceptible mosquitoes in which a number of parameters depend on temperature (*T*) (see Section 4 of the Supporting Information). By assuming fast dynamics of the mosquito (so that levels of infection in the mosquito population quickly equilibrate to the dynamics of infection in the human population), we derive the following expression for the effective transmission rate in the mosquito-human model in terms of the biting rate *a(T)*, probability of human infection given an infectious bite *b(T)*, probability of mosquito infection given biting of an infectious human *pMI(T)*, adult mosquito mortality rate μМ, carrying capacity K of the mosquito population, human population size N, and mosquito demographic function g(T):

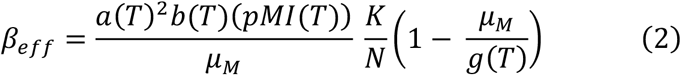

The function g(T) is the product of the eggs laid per female mosquito per gonotrophic cycle, the mosquito egg-to-adult survival probability, and the mosquito egg-adult development rate divided by the adult mosquito mortality rate μМ. The temperature-dependence of these components was borrowed from the literature (47, 48) (see Supporting Information Section 4 for details).

Under the fast dynamics assumption, this effective transmission rate β_eff_ is an implicit representation of the force of infection inflicted on humans by the vectors of the coupled human-vector model. When re-scaled between 0 and 1, β_eff_ corresponds closely with β_MLE,_ the transmission rate from the fitted stochastic SIR cosine model (Panel B of Supplemental Figure S19). This close correspondence indicates that the SIR cosine model is able to capture the shape of the seasonality of DENV1 in Rio de Janeiro.

## Discussion

We developed two lines of evidence regarding the uncertainty and predictability of the time to re-emergence for diseases with low reproductive numbers, on the basis of a seasonally forced SIR model under the ‘well-mixed’ assumption at aggregated, city-wide, scales. We showed with an analytical approach that the time to re-emergence (expressed as the number of “skip” years) was highly sensitive to small changes in *R*_0_ and the fraction of the population still susceptible *s*_*0*_ at the time of prediction (e.g. at the end of the initial outbreak). This sensitivity applies to dengue in Rio de Janeiro where re-emergence times can vary on the order of decades based on literature parameters. This uncertainty contrasts with previous applications of this analytical approach to SIR dynamics in childhood diseases such as measles with much higher *R*_0_ values where accurate predictions of much shorter skip times have been made (29). We also showed with a stochastic SIR model with seasonal transmission fit to DENV1 observed case data for Rio de Janeiro from 1986-1988 that susceptible depletion and replenishment are insufficient to explain dengue re-emergence. The fitted model failed to predict by far the re-emergence of DENV1 in 1990 in terms of either the number of skips expected or the outbreak probability under forward simulation.

Transmission parameters like *R*_0_ are generally defined with respect to a particular model. Given that we aggregated cases at the city level and used a short time series, care should be taken in interpreting parameter values. Nevertheless, fitted transmission parameters correspond well with literature values and exhibit well-defined confidence intervals. Estimates of the reporting rate in particular closely match the 3% value (8) obtained via a serological study conducted during the 1986 invasion (8, 33). Reporting rates during the onset of an epidemic may be much lower in regions that have not recently experienced transmission (33, 49) than in those with re-occurring outbreaks and an established surveillance network. This may explain why serological studies of the 1986 invasion (8, 33) and our results, estimate a lower reporting rate for dengue than studies conducted in later years in Brazil (50). Even though different combinations of the transmission rate and duration of infection can yield the same reproductive number, the parameter estimates that compose *R*_0_ across all models considered in the sensitivity analysis (which take into account those different combinations) are relatively well-defined. These values are also consistent with the effective reproductive number estimated for local dengue epidemics from 2012-2016 (44) and 1996-2014 (45) taking into account differences in serotype circulation and population size during those periods.

More complex model structures are possible and often used for arboviruses that include an explicit representation of the vector. We expect our results to hold as this vector component should largely affect the phase and shape of seasonality in the transmission between human hosts, which we have modeled phenomenologically as a cosine wave. With two typical successive epidemic years from an emergent virus, parameter inference from such short observation period is unlikely to justify a more complex model. Nevertheless, to examine transmission seasonality further, we compared the seasonal *R*_0_ resulting from the fitted model to the seasonal *R*_0_ directly estimated from time series of cases in the literature (44). We also considered the transmission rate experienced by humans in a simple vector-human model forced by the typical seasonality of temperature in Rio de Janeiro. The shape and timing of the vector-human model’s transmission rate was comparable to that of the cosine transmission rate we employed. More complex models that do not assume fast dynamics of infection in the vector relative to epidemic spread would likely exhibit a difference relative to our transmission rate, especially a delayed phase, whose consequences should be examined in future work. We posit that this difference would not influence our results on the predictability and uncertainty of re-emergence, since the values of other parameters (such as the length of infection in humans) can compensate for it.

Factors that could explain the observed rapid re-emergence include inter-annual climate anomalies, antigenic evolution, or micro-scale spatial heterogeneity in transmission intensity and associated susceptible depletion. Larvae washout following flooding coupled with temperature-driven seasonality in transmission could have temporarily halted the invasion in 1988 and delayed the epidemic in 1989. Widespread flooding was reported in February 1988 (51). Large amounts of rainfall washed away mosquito larvae in lab and field studies (52). High rainfall negatively affected dengue transmission in Singapore (53, 54) and India (55). The impact could be compounded in Rio de Janeiro if the high rainfall occurs during the transmission season. If the larvae population has not fully recovered before the start of the off-season, the impact of the rainfall anomaly could extend to the subsequent season.

The large amount of process noise observed in the aggregate model would be consistent with this effect, given that the process noise parameter σ_P_ represents random variation in the transmission rate due to environmental factors. However, the model’s inherent structure limits its ability to take into account flooding events via *σ*_P_, since the magnitude of the process noise does not change between years. Incorporating an inter-annual climate driver could provide more accurate re-emergence predictions. The response to rainfall would be nonlinear: positive at low to moderate levels and negative at higher ones.

Intra-serotype antigenic evolution from 1986-1990 could also facilitate faster re-emergence. Many models focus on inter-serotype variation and assume long-lasting homosubtypic immunity (18, 19, 21). However, the antigenic variation within and across dengue serotypes is comparable (56), and antigenic differences between strains of the same serotype influence overall dengue evolution(57). Sequences associated with case data were unavailable, making direct analysis challenging. We cannot rule out the possibility that genetic differences between the circulating strains enabled re-infection. A future SIRS-type model (Susceptible-Infected-Recovered-Susceptible) could incorporate this intra-serotype antigenic evolution.

Micro-scale spatial heterogeneity in transmission intensity and the effects of human movement between neighborhoods could also explain the rapid re-emergence. Small-scale differences in socioeconomic status and population density between neighborhoods in a large city can result in different relationships between mosquito and human population sizes, resulting in widespread heterogeneity in *R*_0_ across neighborhoods (58). Previous studies of mosquito trap data in the city have demonstrated that neighborhoods with differing socioeconomic characteristics have different vector population patterns (46). In fact, schoolchildren from neighborhoods with divergent socioeconomic characteristics had varying levels of seroconversion during the 1986 invasion (33). Human movement between neighborhoods may also influence transmission within (59) and between (60) those neighborhoods, potentially resulting in non-uniform depletion of susceptible populations between highly connected and isolated areas of a city. Whether arising through the effects of spatial heterogeneity in transmission or intra-city movement, non-uniform levels of herd immunity could enable faster re-emergence.

Our findings reveal the uncertainty of re-emergence predictions with the simplest SIR models, those that would be most useful at times of emergent public health threats. Consideration of the above factors in transmission models whose goal is to inform public health over large regions, and to do so soon after, if not during, an emergent outbreak, is clearly a challenge. For example, coarse resolutions are typically used because of the scales at which the observed cases are reported, the scales at which the climate covariates are available, and the difficulties inherent in incorporating microscale variation including connectivity. Our results should motivate further research into the central question of how we can scale microscale heterogeneity to formulate aggregated models that include it implicitly. It should also motivate the related further understanding of how such microscale heterogeneity influences susceptible depletion and replenishment in particular case studies. From such efforts, we should be able to evaluate whether the increasing availability of high-resolution data makes it feasible to parameterize transmission models at higher resolutions, or to inform new model formulations at coarser resolutions.

The inability of susceptible depletion and replenishment in a simple seasonal SIR formulation at a large, city-wide scale, to explain DENV1 re-emergence has potential implications for other arboviruses. Recent long-term Zika forecasts (31) assume that susceptible depletion and replenishment brought an end to the 2015-2017 epidemics and will determine when re-emergence occurs. DENV1 and Zika share the same vector and invaded a completely susceptible population (not accounting for pre-existing cross-immunity from dengue). If factors absent from the basic model were key drivers of DENV1 inter-annual variability, it would not be unreasonable to infer that similar types of factors could have played a major role in the Zika dynamics observed from 2015-2017. Zika re-emergence could similarly occur much earlier than expected.

With changing temperature patterns due to climate change, cities in Asia, Europe, and the western hemisphere that currently do not have recurrent local transmission may transition in the near future to the kinds of dynamics studied here. Our results suggest that estimates should be interpreted in the context of this sensitivity to small changes in the reporting rate and reproductive number. Factors like variation in reporting rates, micro-scale transmission heterogeneity and inter-annual climate drivers that are often ignored in long-term forecasts may thus become critical in determining re-emergence times. Overall, the large uncertainty in re-mergence times may be unavoidable for these regions. Improved models are needed together with richer data than currently used, to address the question of the relevant spatial scales of susceptible depletion.

## Materials and methods

The derivation of the expression for the number of skip years (Equation 1) is included in Section 1 of the Supporting Information. We fitted a stochastic version of the SIR model to observed monthly case counts in Rio de Janeiro from 1986-1988 to estimate parameters needed to apply this expression, and also to separately predict in parallel the time to re-emergence via numerical simulation Expected re-emergence times were then compared for the two approaches.

### Data Description

We used monthly dengue case estimates in the city of Rio de Janeiro, Brazil from 1986-1990. Cases were reported to the local public health surveillance system (9, 10). The case counts did not contain serotype information, but prior studies indicated that the dengue serotype DENV1 invaded the city of Rio de Janeiro in 1986 (10) and was the dominant serotype in circulation in the state of Rio de Janeiro from 1986-1990 (8) prior to the arrival of DENV2 in 1990. DENV2 did not become dominant until 1991 (9).

### Basic Model Formulation

Because dengue infection confers full immunity to the same serotype, we considered an SIR (Susceptible-Infected-Recovered) model. The deterministic model for the number of individuals in the Susceptible *(S)*, Infected (*I)*, or Recovered (*R)* class is given by the following system of ordinary differential equations:

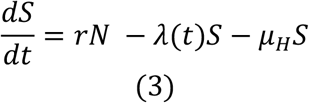

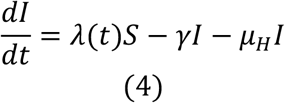

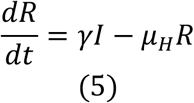

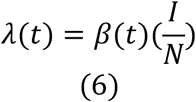

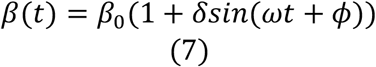

Deaths occur at rate (μ_H_) given by the inverse of the life expectancy of Brazil in 2012 (74.49 years(35)). All individuals are born susceptible. The term *r* represents population growth. The human population growth rate was estimated from census resident population estimates in 1991 (36) and 2000 (37) assuming exponential growth. This rate was used to interpolate the estimated population in 1986 (See Supporting Information Section 2.1.1 for details).

The per capita rate at which susceptible individuals become infected was given by the force of infection λ(t) (Equation 6). Individuals recovered at per-capita rate γ whose inverse is the duration of infection. Estimates of the duration of infection in dengue vary. One analysis estimated that symptoms of dengue infection last 2-7 days following an incubation period of 4-10 days (61, 62). For our analysis, we fixed the recovery rate γ to be 1/17, assuming an exponentially distributed duration of infection with mean of 17 days encapsulating the maximum extent of the combined incubation and symptomatic period in humans. We take into account the possibility that duration of infection could vary by profiling over the duration of infection in the sensitivity analysis. The short duration of the available time series meant that fitting a formal vector model could prove difficult and could require additional assumptions in terms of which parameters could be fitted or fixed from existing formulations in the literature. We therefore used an SIR framework in which the infected stage served as a proxy for the exposed and infected human compartments in a vector model of dengue transmission, and we assumed infection levels in vector rapidly equilibrate with those in humans (as described later when we consider the vector explicitly (See Section 4 of the Supporting Information). A duration of infection was thus chosen that corresponds to the upper bound of the estimated pre-infectious period (4-10 days) and infectious period (2-7 days) in humans (61, 62). We profiled over the duration of infection in the sensitivity analysis to verify that this parameterization is reasonable.

This transmission rate *β(t)* was represented as a cosine function with mean β_0_, (units of contacts per person per day) and seasonal oscillations of amplitude δ (same units as β_0_) and frequency ω, which was assumed to be annual (ω = 2π/365) days^-1^. The annual mean *R*_0_ was thus given by:

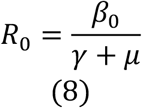

The observed dengue data in Rio de Janeiro consisted of monthly case counts. Serological studies of the DENV1 invasion in Rio de Janeiro also indicated substantial under-reporting (8, 33). Let *C* represent the true number of monthly cases that would be obtained by summing the number of individuals entering the infected class (*I*) over the course of a month. For the purposes of the skip analysis, we assume that a fixed fraction ρ of the true cases *C* are observed, where ρ is the reporting rate.

The stochastic model is an approximation of the deterministic one used for the skip analysis. For simplicity and given the short time interval, we assumed that there was no population growth over the two and half years of the DENV1 invasion (*r =*μ_H_) and that births and deaths occurred at rate μ_H_ = (1/(74.9*365)), which is equal to the inverse of the average life expectancy in Brazil from the 2010 census (35). However, population growth is taken into account when simulating forward in time from the fitted stochastic model. We also assumed that there were no recovered individuals at the start of the epidemic, so all other individuals in the population not initially infected were susceptible. We considered time in units of days and used a time step Δt of 1 day. The stochastic model is a discrete-time model with fixed time step Δt and a discrete state space (i.e. the number of people in each compartment *S, I, R*, and *C*, at any point in time must be integers). The number of individuals who moved from one compartment to another over the course of each day was calculated via Euler simulation from the deterministic equations (See Supporting Information). Demographic stochasticity was then incorporated into the Euler approximations to obtain integer state variable values after each time step. We specifically assumed that the number of individuals making each state transition was drawn from a binomial distribution with exponentially decaying probability (See Supporting Information). Environmental noise (variation in the transmission rate β(t) due to random environmental variation) was captured via multiplicative gamma white noise in the transmission rate as described by (63, 64). On time step size Δ t, we multiplied the transmission rate by ΔГ / Δ t, where ΔГ / Δ t was drawn from a Gamma distribution with mean 1 and variance σ_P_^2^/ Δ t.

The measurement model assumed that the observed number of monthly dengue cases (*Y(t)*) at time *t* were drawn from a negative binomial distribution with mean equal to the true number of monthly cases *C* multiplied by a reporting rate ρ, with dispersion parameter σ_M_. More details of the measurement model can be found in Section 2.4 of the Supporting Information.

### Fitting the stochastic model

We fitted the transmission parameters (β_0_ and δ), reporting rate (ρ), process noise parameter (*σ*_P_), measurement noise parameter (*σ*_M_), and the number of infected individuals at the start of the outbreak in May 1986 (*I*_0_). While the first cases of DENV1 were reported in April 1986, we started the model fitting in May 1986 to avoid complications from changes in the reporting rate as the surveillance system was established during the start of the DENV1 invasion. We used in an interpolated initial population size of 5,281,842 for Rio de Janeiro in May 1986.The model was fit using the mif2 method in the R-package pomp. The model fitting method is described further in the Supporting Information and in (65).

### Calculating expected skips using parameter estimates from stochastic model

Following the completion of the Monte Carlo Profiles, a maximum likelihood estimate (MLE) parameter combination was obtained from the Monte Carlo Profiles of the fitted model by selecting the parameter combination with the highest likelihood across all profiles. The table of MLE parameter values is shown in Supplemental Table ST1. All sets of parameter combinations within 2 log-likelihood units of the maximum likelihood estimate (from all profiles) were used for the expected skip calculation. The reporting rate (ρ), β_0_, and δ value of each parameter combination within 2 log likelihood units of the maximum likelihood estimate were applied to a finer gridded version of the deterministic skip calculation described earlier. A distribution for the number of skips expected in Rio de Janeiro following the DENV1 invasion from 1986-1988 was obtained.

### Stochastic Simulation

We then simulated re-emergence probabilities under the stochastic model. Each parameter combination within 2 log likelihood units of the MLE estimate from the stochastic fit was simulated again without any immigration from 1986 until 1990 but with population growth. During January 1990, “sparks” of infectious individuals were assumed to have arrived in the city at some fixed rate. There were low but non-zero levels of DENV1 incidence from 1988-1989. We chose to wait until January 1990 before introducing new DENV1 infections to be conservative, as this is when an uptick in DENV1 incidence was first observed. Had we introduced sparks earlier in 1988-1989, we would likely have observed even earlier re-emergence times. We explored rates from 5 to 100 infected individuals per day. This process was repeated 100 times, and the probability of an epidemic occurring in 1990 was calculated. An epidemic occurrence in this situation was defined as a net decrease in the susceptible population over the course of the year (after taking into account population growth), to best match the definition of an epidemic used in the skip analysis.

### Sensitivity Analysis

We assessed how parameter estimates of *R*_0_ and ρ may depend on the model formulation by fitting several more complex SIR-type models to the same data using the fitting procedure described in the Methods section: an SIR Spline Model and SEIR Spline Model. As an additional sensitivity analysis, we profiled over the recovery rate for the SIR Cosine Model (Supplemental Figure S9). For details, see the Supporting Information.

### Comparison with Vector Model and literature R_0_

For a full description of the explicit coupled human-mosquito model with compartments for infectious and susceptible mosquitoes and comparison of transmission rates between this model and the simpler seasonally forced SIR, see the Supporting Information.

## Data Availability

Data and code used to fit the stochastic model will be included via a link to a public github repository upon publication along with Supplementary Information.

## Author Contributions

RS, VRA, MP designed the experiments. RS and VRA conducted the experiments. RS and EI developed the stochastic model fitting pipeline. CC provided the data and expertise on dengue in Rio de Janeiro. RS, VRA, and MP drafted the manuscript. All authors contributed to the writing of the manuscript.

## Acknowledgements

The authors would like to thank Aaron King for his advice, and two referees for their insightful comments. This work was completed with resources and support provided by the University of Chicago’s Research Computing Center.

## Data Accessibility

Data and code used to fit the stochastic model is accessible via a public GitHub repository (https://github.com/pascualgroup/JRSI_DENV1_skips_Rio).

## Ethics

This article does not present research with ethical considerations. The authors declare that they have no competing interests.

